# Development and Evaluation of a new Swiss Made SARS-CoV-2 antigen-detecting rapid test

**DOI:** 10.1101/2021.03.25.21252280

**Authors:** Ducrest P.J.

## Abstract

There is an urgent need in rapid diagnostic test (RDT) to detect antigen of SARS-CoV-2 to reduce the spread of COVID-19 outbreak. We have developed a rapid and simple point-of-care lateral flow immunoassay (LFIA) detecting nucleocapsid antigen of SARS-CoV-2 in 10 minutes. The aim of this study is to evaluate the diagnostic performance and analytical sensitivity of this RDT. RT-PCR positive nasopharyngeal swab samples (n=20) for SARS-CoV-2 and 40 negative control samples were studied. Analytical sensitivity was assessed using Gamma-irradiated SARS-CoV-2 and the limit of detection (LOD) was determined at 1.4 × 10^2^ TCID_50_/ml. Overall, RDT diagnostic sensitivity was 90% (95% confidence interval [95%CI]: 67-98%) and specificity 98% (95% CI: 85-100%). The sensitivity was 100% (95% CI: 75-100%) when using only samples with a RT-PCR Cycle threshold lower than 30. This antigen RDT displays a high diagnostic accuracy for SARS-CoV-2 antigen detection in high COVID-19 prevalence settings. Its use could be considered in the absence of routine RT-PCR facilities such in low-income countries.

## Introduction

The COVID-19 pandemic is a challenge for the population, healthcare systems and manufacturer of medical devices. The easy and rapid detection of infected patient is crucial to shorten the spread of the SARS-CoV-2. The actual gold standard for the diagnostic of COVID-19 is the reverse-transcription Polymerase Chain Reaction (RT-PCR) but has certain inconveniences such as the time, laboratory and equipment needed. Such analyses can difficulty be implemented in low-income countries where laboratory or trained staff are missing [1-3].

Rapid antigen tests are lateral flow immunoassay with a visual (gold nanoparticles) or fluorescent detection. Since the beginning of the COVID-19 outbreak, thousands of rapid tests have been listed on the COVID-19 Testing pipeline of the Find Foundation (Geneva, Switzerland). Most of the test are produced in Asia.

Recent studies highlighted a lower sensitivity compared to RT-PCR [4, 5]. However, the most common rapid antigen test such as Panbio (Abbott), Standard Q (SD Biosensor, Roche Diagnostics) and BD Veritor (Beckton & Dickinson) are the most studied and published tests. Based on a meta-analysis of available performances data, the overall sensitivity for Panbio (Abbott) is 79% and the specificity is 99% (n=10936). For the Standard Q (SD Biosensor, Roche Diagnostics), the overall sensitivity is 85% and specificity 99% (n=2892). Finally, the BD Veritor (Beckton & Dickinson) has an overall sensitivity of 91% and a specificity of 99% (n=964) [6-9]. It has been noted that the sensitivity increases with high viral load (RT-PCR Cycle threshold < 30). The World Health Organization (WHO) published a target product profile with the minimal and desirable analytical and clinical performances (TPP WHO) [10]. The WHO recommended that an Ag-RDT test must have a sensitivity of at least 80% and a sensitivity of at least 97%, based on the gold-standard RT-PCR test. The analytical performance must be at least 10^6^ genomic copies/ml (CT 25-30).

A patient with a positive Ag-RDT result within 5 days after symptom onset can be considered to have a SARS-CoV-2 infection, because these individuals are more likely to have high viral loads [11]. However, a negative result must be interpreted with caution due to reduced sensitivity with low viral load and a confirmation test (RT-PCR) is recommended [10].

Here we have developed and evaluated a new rapid antigen-detecting test following the last recommendation of the WHO. Both analytical performance and clinical performances have been evaluated.

## Material & Methods

### Study population and swab sample collection

Anonymized leftovers of frozen Nasopharyngeal swab specimen, discharged in medium Tris EDTA 1X, supplied by INO Specimens BioBank, ISB (Clermont-Ferrand, France), were used for the evaluation. A total of 60 samples, including 40 negative and 20 positive samples were all evaluated with CE-IVD & EUA-FDA reference RT-PCR Nucleic Acid Diagnostic Kit (Sansure Biotech Inc, Changsha, China) on the QuantStudio 7 Pro Real-Time PCR Systems Instrument (Termofisher Scientific, Waltham, USA). The positive samples were classified according to the Cycle Threshold (CT) for both probe *orf1 ab* and *N gene*. The cut-off values for RT-PCR CT were < 39.00 for positive samples and > 42.00 for negative samples.

### Inactivated SARS-CoV-2 and recombinant proteins

SARS-Related Coronavirus 2, Isolate USA-WA1/2020, Gamma-Irradiated and Heat-inactivated were supplied by BEI Resources. Recombinant SARS-CoV-2 Nucleocapsid protein was supplied by GenScript Biotech (Netherlands) B.V. (Leiden, NL). Tenfold dilution was prepared in the running buffer of the test and evaluated in triplicates. The limit of detection is defined as the lowest concentration were the signal intensity is egal or greater than 2 Rann score value (BBI, UK).

### Antigen Rapid test

The Antigen-detecting rapid test COVIDia-Antigen was developed and manufactured in Switzerland by GaDia SA (Monthey, Switzerland). This test detects the nucleocapsid antigen in nasopharyngeal and throat swabs. The test has two pre-coated lines on a nitrocellulose membrane, one Control line (C) and one Test line (T). The test line is composed of rabbit monoclonal anti-nucleocapsid antibody. The gold nanoparticles (Nanocomposix, San Diego, USA) are coated with rabbit anti-Nucleocapsid antibodies. The control line (C) is composed of a goat anti-rabbit antibody (Jackson ImmunoResearch Europe, Ely, UK). When a positive sample flows through the test membrane, the anti-SARS-CoV-2 antibodies present on the membrane (Test line) capture the antigen and form a colored conjugate complex on the test line. The control line captures the excess of gold conjugate nanoparticles and forms a red line.

### Sample Testing

The pouched test was opened immediately before use. To perform the test, 60 µL of defrosted Nasopharyngeal swab sample was diluted in 60 µL of running buffer according to the manufacturer instructions. Then, 100 µL were added to the sample hole and the test was read after 15 minutes. For the limit of detection using inactivated SARS-CoV-2, the sample was directly diluted in running buffer and 100 µL were added to the test as describe above.

### Statistics

Vassarstats online tool (www.vassarstats.net) was used to calculate sensitivity (SE), specificity (SP), positive and negative predictive values (PPV, NPV), 95% confidence intervals, median, and Interquartile range (IQR); while significance (p-values) was calculated using Mann-Whitney U test for categorical variables. Statistical significance was defined as p < 0.05.

## Results

### Baseline characteristics

The baseline demographic characteristics of patients’ samples were as follows: the 20 RT-PCR confirmed COVID-19 samples from patients were older (median 63 years old, IQR 44-80.75) compared to the healthy plasma samples patients (n=40) (median 51 years old, IQR 39-70.75; p<0.05). The proportion of females in COVID-19 cohort was 75% (n=15) and 43% (n=17) in negative control group. The median CT value for *N gene* probe is 25.03 (IQR 20.1-30.6) and for *orf1 ab* probe is 26.8 (IQR 21.5-30.2).

### Specificity of Antigen RDT on negative control group

Antigen RDT diagnostic specificity (percent negative agreement, PNA) against negative control group (n=40) is shown in Table 1. Antigen RDT revealed concordant results in 39 of 40 samples. Only one discordant result showed a low positive signal, while being negative (false-positive). These resulted in an Antigen RDT specificity (SP) of 97.5% (95% CI: 85.3-99.9%).

**Table 1:** Diagnostic performance of antigen RDT

|  |  | RT-PCR |  |  |
| --- | --- | --- | --- | --- |
|  |  | Positive | Negative | Total |
| COVIDia-Antigen | Positive | 18 | 1 | 19 |
|  | Negative | 2 | 39 | 41 |
|  | Total | 20 | 40 | 60 |
| Sensitivity |  | 90.0% (CI<sub>95%</sub>: 66.9-98.2%) |  |  |
| Specificity |  | 97.5% (CI<sub>95%</sub>: 85.3-99.9%) |  |  |
| PPV |  | 94.7% (CI<sub>95%</sub>: 71.9-99.7%) |  |  |
| NPV |  | 95.1% (CI<sub>95%</sub>: 82.2-99.2%) |  |  |

### Sensitivity of Antigen RDT against RT-PCR confirmed COVID-19 samples

Antigen RDT diagnostic sensitivity (percent positive agreement, PPA) against RT-PCR confirmed samples (n=20) is shown in Table 1. Antigen RDT revealed concordant results in 18 of 20 samples. Only 2 samples, with a CT value >30 were negative (false-negative). These results indicate an Antigen RDT sensitivity (Se) of 90.0% (95% CI: 66.9-98.2%). When considering samples with a CT <30, the sensitivity is 100% (95% CI: 74.7-100%). The table 2 indicate the Sensitivity or PPA compared to CT values of samples.

**Table 2:** Percent positive agreement of the antigen RDT at different RT-PCR CT values.

| CT values | Percent positive agreement (CI95%) |
| --- | --- |
| <20 | 100% (46.3-100%) |
| 20-25 | 100% (46.3-100%) |
| 25-30 | 100% (46.3-100%) |
| <30 | 100% (74.7-100%) |
| >30 | 60.0% (17.0-92.7%) |
| overall | 90.0% (66.9-98.2%) |

The two false-negative samples have a CT value (N gene) of respectively 31.05 and 35.01 visible on red on the figure 1. The signal intensity was inversely proportional to the RT-PCR CT value. The smaller the CT value, the highest the signal intensity. Based on the regression done on the 20 positive samples (figure 1), R-squared was 0.82 and the LOD with a Rann value of 2 can be estimated at a RT-PCR CT value of 31.5.

**Figure 1:**
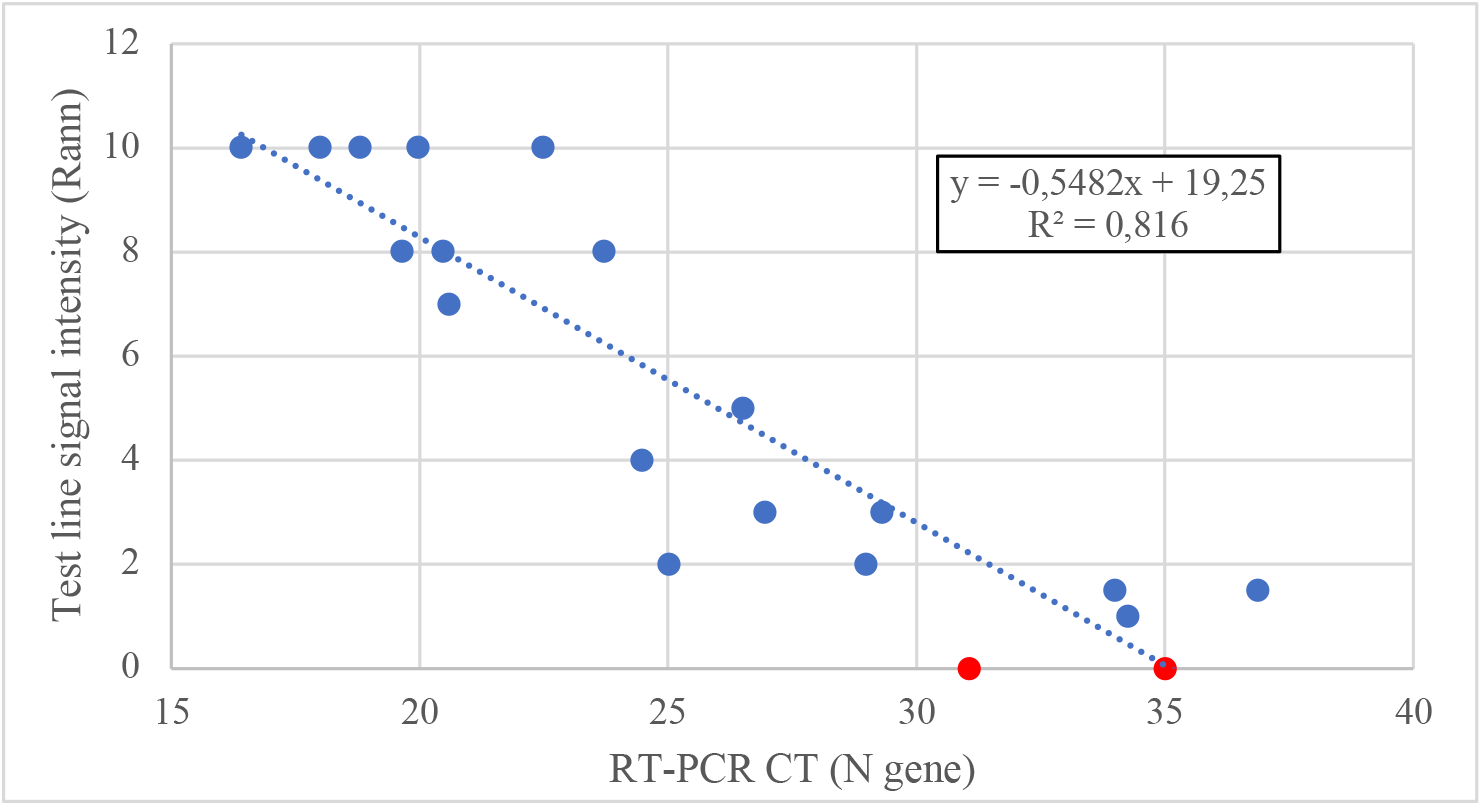
Test line intensity of the antigen RDT compared to the CT value of RT-PCR (N gene). Blue dots represent true-positive samples and red dots false-negative results

### Analytical Sensitivity of Antigen RDT

The analytical sensitivity of the Rapid diagnostic test was evaluated using Gamma-irradiated SARS-CoV-2 (Isolate USA-WA1/2020) from BEI Resources. The irradiated SARS-CoV-2 was diluted in running buffer and tested with rapid test 5 times. The LOD was defined as a signal intensity of 2 Rann score value on all 5 replicates. The table below (table 3) give the results of the LOD evaluation. The LOD of the Rapid test was defined at 1.4 × 10^2^ TCID_50_/ml, corresponding to 7.9 × 10^4^ genome equivalent/ml.

**Table 3:** Limit of detection of the antigen RDT using Gamma-irradiated SARS-CoV-2

| SARS-CoV-2 strain | Gamma-Irradiated SARS-CoV-2, Isolate USA-WA1/2020<br>[BEI Resource, ref. NR-52287, Lot. 70039067] |  |  |  |  |  |
| --- | --- | --- | --- | --- | --- | --- |
| Stock titer (TCID<sub>50</sub>/ml) | $2.8 \times 10^6$ | | | | | |
| Stock titer (genome equivalent/ml) | $1.57 \times 10^9$ | | | | | |
| Dilution | 1/10 | 1/100 | 1/1000 | 1/10000 | 1/20000 | 1/40000 |
| Concentration in dilution (TCID<sub>50</sub>/ml) | $2.8 \times 10^5$ | $2.8 \times 10^4$ | $2.8 \times 10^3$ | $2.8 \times 10^2$ | **$1.4 \times 10^2$** | $7.0 \times 10^1$ |
| Concentration in dilution (genome equivalent/ml) | $1.6 \times 10^8$ | $1.6 \times 10^7$ | $1.6 \times 10^6$ | $1.6 \times 10^5$ | **$7.9 \times 10^4$** | $3.9 \times 10^4$ |
| Signal intensity (Rann) | 10 | 6 | 4 | 3 | **2** | 1 |
| Replicates | 100% (5/5) | 100% (5/5) | 100% (5/5) | 100% (5/5) | **100% (5/5)** | 60% (3/5) |

## Discussion

The key finding of the present evaluation study, using an unmatched case-control study including 33.3% of negative control samples, is that the diagnostic accuracy of antigen RDT on frozen nasopharyngeal swabs when using RT-PCR as reference method, displayed a Percent positive agreement (Sensitivity) of 90.0%, a SP of 97.5%, a PPV of 94.7% and a NPV of 95.1%. These results are considered as desirable for the sensitivity and acceptable for the specificity according to the Target Product Profile of the WHO.

The two false-negative results obtained with this RDT had a RT-PCR CT value bigger than 30, such samples have less viral load. The analytical sensitivity of this RDT was defined at 1.4 × 10^2^ TCID_50_/ml, corresponding to 7.9 × 10^4^ genomic copies/ml (Table 3). The analytical sensitivity based on RT-PCR CT value, was also extrapolated using the 20 positive samples (figure 1) and was defined at a CT value of 31.5. The TPP from WHO defined an analytical sensitivity of at least 30 cycle threshold (CT) values [10]. A Point-of-Care (POC) test that can consistently detects the most infectious patients, with high viral load of at least 10^6^ genomic copies/ml, in order to interrupt transmission. This RDT fulfills the TPP of the WHO in term of sensitivity and specificity. The diagnostic sensitivity is comparable to the SD Biosensor and Abbott Panbio rapid antigen tests [7, 8].

Such performances indicate that this RDT could be fit for purpose in clinical settings where a high prevalence of COVID-19 prevails, especially in situations where RT-PCR are not available or cannot be reliably used. Diagnostic performances in low prevalence populations still needs to be determined and larger populations need to be tested. In addition, it is clear that antigen assay for SARS-CoV-2 need to be performed in the first week after symptoms onset [6, 12]. Finally, here we used frozen nasopharyngeal swab specimens and the test was performed in a laboratory environment; we may expect different results in real-life at patients’ bed and using fresh nasopharyngeal swab.

In addition, the test provided clear results, without indeterminate or invalid results (no Control line). There are several limitations to this study. First, we present here the results of a method evaluation study and not a seroprevalence study. Therefore, the PPV obtained here (based on a 33.3% proportion of cases defined as laboratory confirmed SARS-CoV-2 by RT-PCR) will probably be lower in a low prevalence setting. Another limitation of this validation study lies in its limited sample size leading to broad 95% confidence intervals, requiring confirmation of these data at a larger scale. Finally, our present conclusions only apply to this RDT, and must not be applied to any other RDTs currently available.

In conclusion, this RDT is not meant to replace a SARS-CoV-2 RT-PCR but could be a reliable option for quickly assessing the presence of SARS-CoV-2 antigen in moderate to high COVID-19 prevalence settings and when high viral loads are expected, especially in situations where a rapid result is needed or where RT-PCR is not available. Further investigations in low prevalence situations and using capillary blood are necessary.

## Data Availability

Data are available in Supplementary material

