## Supplementary Data for "Development and Evaluation of a new Swiss Made SARS-CoV-2 antigen-detecting rapid test"

*Supplementary Material: Antigen rapid test clinical evaluation results*

| | | | CT RT-PCR Sansure Biotech (CE-IVD & EUA-FDA) | | COVIDia-Antigen Result | |
|---|---|---|---|---|---|---|
| **Sample #** | **Age Range** | **Gender** | **Probe orf1 ab** | **Probe N gene** | **Rann Score** | **POS/NEG** |
| 1 | >80 | M | 18.60 | 16.41 | 10 | POS |
| 2 | >80 | F | 19.05 | 17.99 | 10 | POS |
| 3 | >80 | M | 21.30 | 18.81 | 10 | POS |
| 4 | 30-40 | F | 20.76 | 19.66 | 8 | POS |
| 5 | 50-60 | F | 22.95 | 19.97 | 10 | POS |
| 6 | 40-50 | F | 21.29 | 20.47 | 8 | POS |
| 7 | 40-50 | F | 22.13 | 20.60 | 7 | POS |
| 8 | 70-80 | F | 23.01 | 22.48 | 10 | POS |
| 9 | 60-70 | M | 24.95 | 23.71 | 8 | POS |
| 10 | >80 | F | 26.80 | 24.47 | 4 | POS |
| 11 | >80 | F | 26.68 | 25.03 | 2 | LOW POS |
| 12 | 70-80 | F | 27.25 | 26.52 | 5 | POS |
| 13 | 20-30 | M | 27.45 | 26.96 | 3 | POS |
| 14 | 40-50 | F | 29.12 | 28.98 | 2 | LOW POS |
| 15 | 60-70 | F | 30.53 | 29.30 | 3 | POS |
| 16 | 50-60 | F | 28.48 | 31.05 | 0 | NEG |
| 17 | 50-60 | F | 34.97 | 33.99 | 1,5 | LOW POS |
| 18 | >80 | F | 34.05 | 34.26 | 1 | LOW POS |
| 19 | 40-50 | F | 37.15 | 35.01 | 0 | NEG |
| 20 | 70-80 | M | 37.14 | 36.88 | 1,5 | LOW POS |
| **NEG** | | | | | | |
| 21 | 70-80 | M | NA | NA | 0 | NEG |
| 22 | 70-80 | F | NA | NA | 0.5 | NEG |
| 23 | 60-70 | F | NA | NA | 0 | NEG |
| 24 | 20-30 | M | NA | NA | 0 | NEG |
| 25 | 30-40 | F | NA | NA | 0 | NEG |
| 26 | 40-50 | F | NA | NA | 0 | NEG |
| 27 | 40-50 | M | NA | NA | 0 | NEG |
| 28 | 40-50 | M | NA | NA | 0 | NEG |
| 29 | 50-60 | F | NA | NA | 0 | NEG |
| 30 | 60-70 | M | NA | NA | 0 | NEG |

| | | | | | | |
|---|---|---|---|---|---|---|
| 31 | <20 | F | NA | NA | 0 | NEG |
| 32 | 20-30 | F | NA | NA | 0 | NEG |
| 33 | 40-50 | M | NA | NA | 0 | NEG |
| 34 | 40-50 | F | NA | NA | 0 | NEG |
| 35 | >80 | M | NA | NA | 0 | NEG |
| 36 | 30-40 | M | NA | NA | 0 | NEG |
| 37 | <20 | M | NA | NA | 0 | NEG |
| 38 | >80 | M | NA | NA | 0 | NEG |
| 39 | 30-40 | M | NA | NA | 0 | NEG |
| 40 | 70-80 | M | NA | NA | 0 | NEG |
| 41 | 30-40 | F | NA | NA | 0 | NEG |
| 42 | 30-40 | F | NA | NA | 0 | NEG |
| 43 | 70-80 | F | NA | NA | 0 | NEG |
| 44 | 50-60 | F | NA | NA | 0 | NEG |
| 45 | 60-70 | M | NA | NA | 3 | POS |
| 46 | 40-50 | F | NA | NA | 0 | NEG |
| 47 | 60-70 | M | NA | NA | 0 | NEG |
| 48 | 40-50 | F | NA | NA | 0 | NEG |
| 49 | 50-60 | M | NA | NA | 0 | NEG |
| 50 | >80 | F | NA | NA | 0 | NEG |
| 51 | 40-50 | F | NA | NA | 0 | NEG |
| 52 | 40-50 | F | NA | NA | 0 | NEG |
| 53 | 50-60 | M | NA | NA | 0 | NEG |
| 54 | 20-30 | M | NA | NA | 0 | NEG |
| 55 | 30-40 | M | NA | NA | 0 | NEG |
| 56 | 70-80 | M | NA | NA | 0 | NEG |
| 57 | 70-80 | M | NA | NA | 0 | NEG |
| 58 | 50-60 | F | NA | NA | 0 | NEG |
| 59 | 20-30 | M | NA | NA | 0 | NEG |
| 60 | 60-70 | M | NA | NA | 0 | NEG |
